# Impact of Pacing Positions on Dyssynchrony: Insights from Virtual Body Surface Mapping

**DOI:** 10.1101/2025.08.26.25334474

**Authors:** Alexander Langanke

## Abstract

**Background:** Right ventricular (RV) endocardial pacing is a standard therapy for bradycardia, but it can lead to left ventricular (LV) dysfunction due to electrical dyssynchrony. While conduction system pacing has emerged as a promising alternative, traditional RV pacing remains common, making the optimization of RV lead position clinically relevant. This study aimed to determine whether dyssynchrony metrics derived from a new four-electrode virtual body surface mapping (VBSM) system could be used to identify optimal pacing sites.

**Methods:** We retrospectively analyzed data from 65 patients with RV pacemakers or ICDs. Using imaging from electronic medical records (EMRs), RV lead positions were mapped to 11 of 18 defined anatomical segments. We obtained measurements for three dyssynchrony parameters (standard deviation of activation times [SDAT], QRS duration [QRSd], and QRSarea) from a VBSM system. A point-biserial correlation was used to evaluate the relationship between these parameters and lead position.

**Results:** Septal positions consistently demonstrated lower SDAT values compared to free wall positions (mean SDAT of 23.64 ms vs. 34.03 ms). SDAT showed a strong positive correlation with non-septal positions (rpb = 0.67, p<0.01), indicating that higher SDAT values are strongly associated with free wall pacing. This association was less marked for QRSd and absent for QRSarea. Our findings showed a weak correlation between dyssynchrony parameter rankings and the hemodynamic performance rankings reported in external literature.

**Conclusion:** SDAT is a valuable parameter for distinguishing between septal and non-septal right ventricular pacing positions. These findings underscore the potential of VBSM as a non-invasive tool for guiding electrode placement to minimize electrical dyssynchrony and potentially optimize hemodynamic outcomes. However, the retrospective nature of the study, small sample size, and reliance on literature-derived hemodynamic data are key limitations. Future direct hemodynamic studies are essential to validate these observations.

## Introduction

Since its introduction, right ventricular (RV) endocardial pacing has become a standard therapy for treating bradycardia, and it is now widely adopted in interventional cardiology departments globally. Pacemaker therapy can be lifesaving in many cases of high-grade atrioventricular (AV) block and significantly enhance the quality of life in other forms of bradycardia.[1]

Despite its benefits, RV pacing is linked to increased left ventricular (LV) dysfunction due to electrical dyssynchrony, similar to left bundle branch block (LBBB).[2,3] Not all patients respond equally negatively to RV pacing, possibly due to differences in baseline LV function or RV electrode position.

While conduction system pacing (CSP) has emerged as a promising alternative, traditional RV pacing remains a common and necessary treatment. Therefore, optimizing the RV electrode position to minimize pacing-induced dyssynchrony remains clinically relevant.

Previous research by Vancura et al. demonstrated that different RV pacing positions resulted in measurable differences in acute LV hemodynamics, with septal pacing showing more favorable outcomes compared to non-septal pacing.[4] In observational studies, septal pacing is associated with a lower incidence of heart failure events.[5] Differentiating between septal and non-septal electrode placements can be complex when relying solely on fluoroscopy, often leading to misidentification.[6]

Body surface mapping is a technique employing vast numbers of electrodes distributed across the thorax using a belt or a vest used to capture a more holistic view of the electrical activity of the heart compared to the 12-lead ecg or even 1-lead ecg often found in operating rooms.[7–10] This allows for better characterization of electrical dyssychrony which is the basis of LV dysfunction in LBBB and RV pacing.[8–10]

Body surface mapping (BSM) is a technique that provides a more holistic view of the heart’s electrical activity than a 12-lead ECG, allowing for better characterization of electrical dyssynchrony. Research has indicated that variations in BSM-derived dyssynchrony metrics, such as the standard deviation of activation times (SDAT) and QRSarea, correlate with changes in LV hemodynamics. [[10–12]

Body surface mapping mainly suffers from its cumbersome use and, depending on the technology used, interference with sterile field during surgery.[13]

This study’s objectives are threefold. We aimed:

1. To determine which of the three measured dyssynchrony parameters (SDAT, QRS duration (QRSd), and QRSarea) best differentiate between different RV lead positions.
2. To assess whether certain RV lead sites, particularly septal positions, correlate with more favorable electrical profiles, and to compare these findings with the hemodynamic data reported by Vancura et al..
3. To explore the potential of a new, four-electrode-based virtual body surface mapping (VBSM) system as a non-invasive, intraoperative tool to guide RV lead placement and potentially optimize patient outcomes. We hypothesized that VBSM-based dyssynchrony measurements would correlate with different RV electrode positions and that hemodynamically optimal positions would correspond with lower dyssynchrony metrics.

## Methods

We retrospectively investigated a group of consecutive patients that visited our outpatient clinic for routine pacemaker and ICD (implantable cardioverter-defibrillator) follow-up for which our electronic medical records (EMR) contained imaging information records (computed tomography (CT) scans, echocardiography, chest radiography) that allowed us to identify the position of the RV electrode within the right ventricle.

For comparison purposes, we adopted Vancura et al.’s definition of 18 anatomical segments of the right ventricle (RV), segmenting the RV horizontally into basal, mid, and apical portions; vertically into inferior, mid, and superior portions; and frontally into septum and free wall. Electrode positions were mapped accordingly. If localization was uncertain based on the available data, the corresponding fields were left blank. For instance, if imaging clearly indicated superior and basal positions but lacked confirmation regarding septum placement, only the superior and basal position data were recorded. Due to the retrospective nature of our study, we could only analyze electrode positions that were present in our patient cohort, resulting in data for 11 of the 18 anatomical segments defined by Vancura et al.

During follow-up dyssynchrony (SDAT, QRSarea) and QRSd measurements of RV paced beats were obtained using a new vectorcardiography based virtual body surface mapping (VBSM) system, CardioSecur Pace by Personal MedSystems. This system uses a 4-electrode derived vector loop to calculate 14,000 virtual unipolar leads, generating beat-to-beat activation time maps to calculate SDAT. This iOS based lightweight system can be used intraoperatively without interfering with the sterile field or fluoroscopy and combines the strengths of 12-lead ecg, vectorcardiography and body-surface mapping. Activation time was defined as time from pacing stimulus to minimum of the first derivative of the QRS waveform also known as the intrinsicoid deflection time. The system also calculated vectorcardiography based parameters such as QRSArea. QRSd was also automatically measured.

Vancura et al. employed systolic index as a hemodynamic parameter, and ejection fraction was not measured. Consequently, our dyssynchrony measurements were compared with the systolic index used by Vancura et al.

No CSP leads were involved in the study. For CRT devices, right ventricular-only pacing was utilized during dyssynchrony measurements. The study included patients with single-chamber leadless pacing systems. All patients were paced in VVI (ventricular paced, ventricular sensed, inhibited) mode at a sufficient heart rate to minimize the risk of fusion beats affecting the measurements.

All patients provided written informed consent. Patient characteristics such as age and sex were not studied. Ethics board approval was given before study initiation.

Data acquisition was performed using Microsoft 365 Excel. Data analysis was conducted using GNU PSPP version 2.0.1. To evaluate the relationship between the dyssynchrony parameters (SDAT, QRSd, and QRSarea) and the dichotomous RV electrode positions (e.g., septal vs. non-septal, basal-apical, and craniocaudal), a point-biserial correlation was used. A p-value of less than 0.05 was considered statistically significant.

## Results

A total of 65 patients were included in the study. Of these 38 had complete localization data. The other 27 patients had at least one localization datapoint. Patients were only included in an analysis if there was data concerning the measurement being analyzed.

Table 1 shows mean dyssynchrony parameters for each segment. In general, septal positions show lower SDAT than free wall positions (septal SDAT mean 23,64ms ±2,64ms; free wall SDAT mean 34,03ms ±6,48ms) (Fig. 1). This association is less marked for QRSd (septal QRSd mean 144,13ms ±18,5ms; free wall QRSd mean 159,37ms ±18,95ms)(Fig. 2) and even less for QRSarea (septal QRSarea mean 27,64ms ±13,51ms; free wall QRSarea mean 30,57ms ±21ms) (Fig. 3).

**Table 1.** Dyssynchrony metrics by segment.

| Segment Number | | Mean SDAT (+-SD) ms | Mean QRSd (+-SD) ms | Mean QRS Area (+-SD) μVs |
| --- | --- | --- | --- | --- |
| 1 | Septal-apical-superior | - | - | - |
| 2 | Septal-apical-mid | - | - | - |
| 3 | Septal-apical-inferior | 24.5 (2.12) | 180 (28.28) | 37.50 (2.12) |
| 4 | Septal-mid-superior | - | - | - |
| 5 | Septal-mid-mid | 21.83 (2.86) | 134.67 (7.45) | 24.40 (20.08) |
| 6 | Septal-mid-inferior | 25 (1.41) | 134 (8.49) | 27.50 (4.95) |
| 7 | Septal-basal-superior | 26 (1.22) | 146 (12.57) | 29.50 (10.88) |
| 8 | Septal-basal-mid | - | - | - |
| 9 | Septal-basal-inferior | - | - | - |
| 10 | FW-apical-superior | 39.50 (14.85) | 158 (19.8) | 24 (14.14) |
| 11 | FW-apical-mid | 34.33 (4.51) | 165.33 (9.5) | 19 (1.41) |
| 12 | FW-apical-inferior | 34.50 (7.9) | 163.25 (18.61) | 38.75 (43.65) |
| 13 | FW-mid-superior | - | - | - |
| 14 | FW-mid-mid | 36.17 (3.31) | 146.86 (13.8) | 30 (19.82) |
| 15 | FW-mid-inferior | 31.75 (6.7) | 168.5 (18.93) | 30.5 (13.92) |
| 16 | FW-basal-superior | 35.5 (0.71) | 158 (19.8) | 23 (1.41) |
| 17 | FW-basal-mid | - | - | - |
| 18 | FW-basal-inferior | 23.5 (9.19) | 168 (16.97) | 49 (39.6) |

Comparison of dyssynchrony metrics by segment. FW = Free Wall of the right ventricle

**Figure 1.**
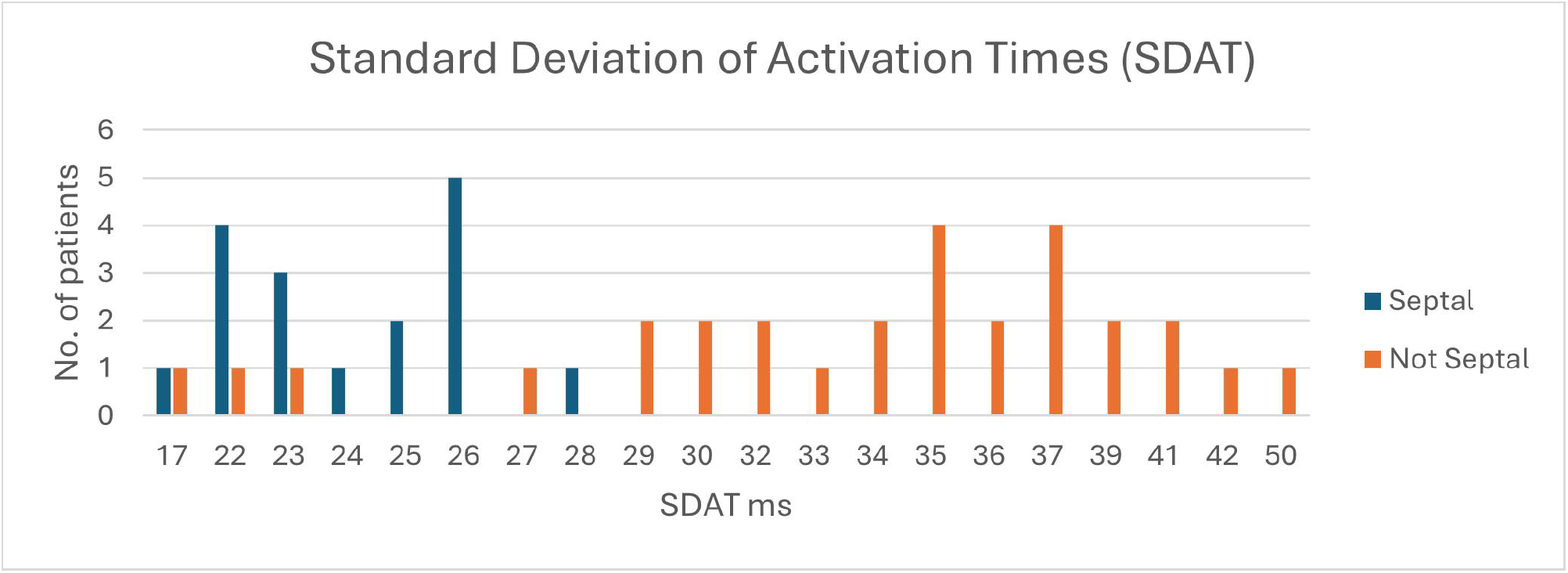
A histogram depicting the distribution of standard deviation of activation time (SDAT) values for septal and non-septal right ventricular (RV) electrode positions.

**Figure 2.**
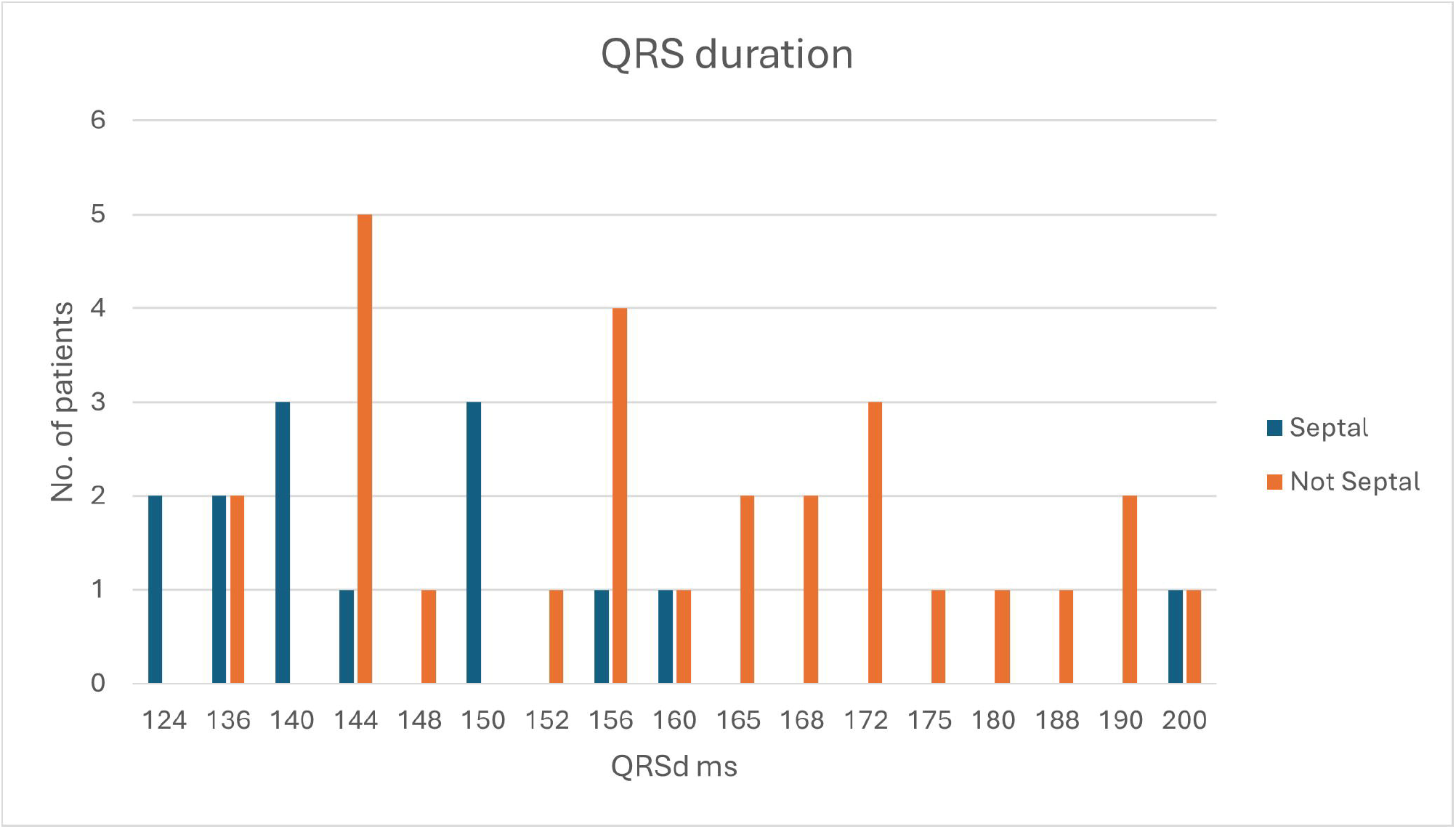
Histogram depicting the distribution of QRS duration measurements for septal and non-septal right ventricular (RV) electrode placements.

**Figure 3.**
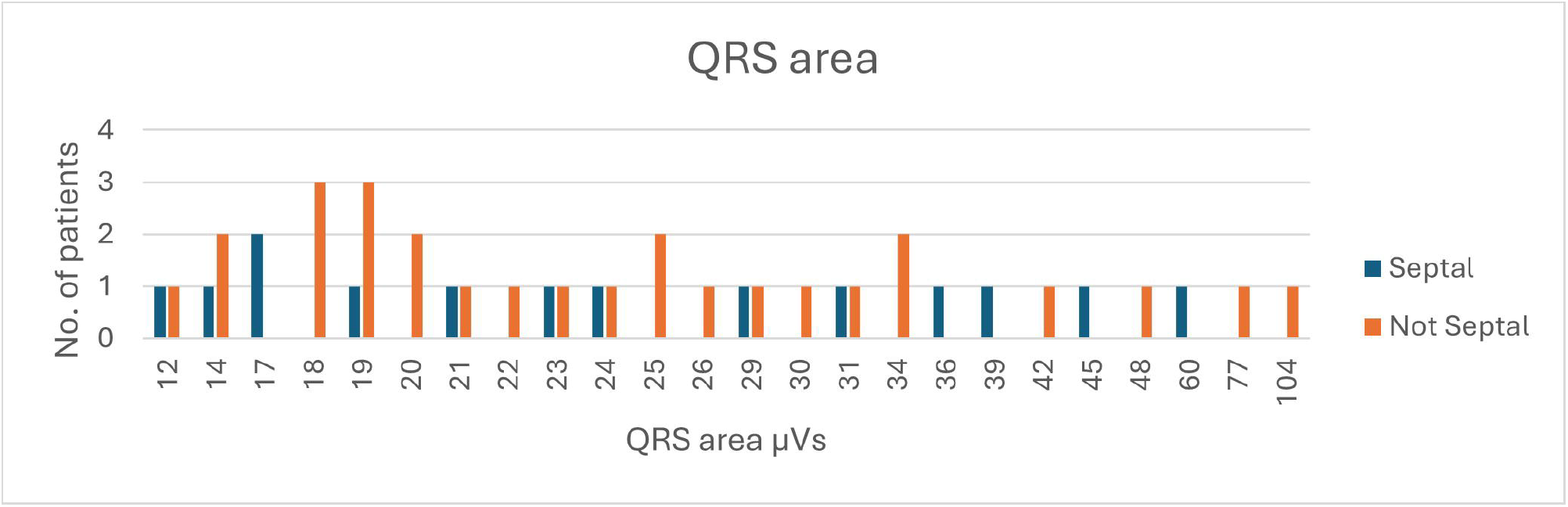
Histogram showing QRS area distribution for septal versus non-septal RV electrode placements.

Table 2 shows the point-biserial correlation coefficients for dyssynchrony parameters in distinguishing between septal and non-septal, craniocaudal, and basal-apical orientations. SDAT showed a strong positive correlation (r_pb = 0.67, p < 0.01) with non-septal positions, indicating that higher SDAT values are strongly associated with a non-septal (free wall) lead placement. QRSd showed a weak but significant correlation for septal-free wall (r_pb = 0.37, p = 0.01) and basal-apical orientations (r_pb = 0.28, p = 0.04). No other significant correlations were observed.

**Table 2.** Dyssynchrony metric correlation.

|  | SDAT | QRSd | QRS Area |
| --- | --- | --- | --- |
| RVOT-non RVOT | -0.065 (p=0.82) | 0.48 (p=0.07) | -0.3 (p=0.28) |
| Septal-non septal | 0.67 (p=0.00) | 0.37 (p=0.01) | 0.08 (p=0.64) |
| caudo-cranial | 0.16 (p=0.3) | -0.23 (p=0.11) | 0.07 (p=0.66) |
| basal-apical | 0.15 (p=0.29) | 0.28 (p=0.04) | -0.12 (p=0.4) |

Comparison of correlation coefficients by spatial orientation of the lead.

Table 3 shows the segments we measured ranked by dyssynchrony metric compared to systolic index in Vancura et al.[4] Since the patients in our study did not have electrodes in all positions Vancura et al. studied, we can only report dyssynchrony measurements for 11 of the 18 positions.

**Table 3.** Comparison of pacing locations by rank.

| Rank # | Vancura et al. syst. Index | Vancura et al. syst. Index (only measured segments) | SDAT | QRSd | QRS Area |
| --- | --- | --- | --- | --- | --- |
| 1 | 8 | 11 | 5 | 6 | 11 |
| 2 | 1 | 5 | 18 | 5 | 16 |
| 3 | 11 | 7 | 3 | 7 | 10 |
| 4 | 5 | 16 | 6 | 14 | 5 |
| 5 | 4 | 3 | 7 | 10 + 16 | 6 |
| 6 | 7 | 14 | 15 | 10 + 16 | 7 |
| 7 | 17 | 12 | 11 | 11 | 14 |
| 8 | 13 | 15 | 12 | 12 | 15 |
| 9 | 2 | 18 | 16 | 18 | 3 |
| 10 | 9 | 6 | 14 | 15 | 12 |
| 11 | 16 | 10 | 10 | 3 | 18 |
| 12 | 3 |  |  |  |  |
| 13 | 14 |  |  |  |  |
| 14 | 12 |  |  |  |  |
| 15 | 15 |  |  |  |  |
| 16 | 18 |  |  |  |  |
| 17 | 6 |  |  |  |  |
| 18 | 10 |  |  |  |  |

## Discussion

This study aimed to investigate whether the hemodynamic effects of different right ventricular (RV) pacing locations could be inferred from vectorcardiographically derived dyssynchrony parameters. The goal was to lay the groundwork for future hemodynamic studies and explore the potential of noninvasive methods to assess the impact of pacing sites during implantation.

Our findings showed only weak correlations between dyssynchrony parameter rankings and the hemodynamic performance rankings reported in Vancura et al. While three out of five segments in our study aligned with the top five segments from the ground truth data for each dyssynchrony metric, significant deviations were observed. Notably, septal positions demonstrated consistently lower SDAT values compared to free wall positions, reinforcing the established notion that septal pacing confers hemodynamic advantages. This finding aligns with existing literature, suggesting that reduced electrical dyssynchrony correlates with improved hemodynamic outcomes.[14]

Interestingly, the trends highlighted in Vancura et al., such as superior positions correlating with better hemodynamic outcomes irrespective of septal or free wall placement, could not be replicated in our study. Among the evaluated dyssynchrony metrics, SDAT proved to be more effective than both QRSd and QRSarea in identifying optimal pacing sites. For instance, only four patients with electrodes positioned in the free wall recorded an SDAT below 30 ms, and only one free wall segment showed a mean SDAT under 30 ms. In contrast, none of the septal segments exceeded this threshold (Fig. 1). This suggests that SDAT could be a dependable indicator for septal positioning and might offer significant clinical value as a cutoff parameter during implantation procedures. Previous research by Ghossein et. al and Okafor et. al indicated that QRSarea could serve as a marker of electrical dyssynchrony, assisting in CRT programming and candidate selection.[15,16] We anticipated a higher correlation, similar to what we observed with SDAT for QRSarea. A plausible explanation for this discrepancy is that QRSarea depends on R-wave magnitude and thus partially on body mass and LV mass. There may have been considerable differences in patient physiognomy in this respect, although this remains speculative since these parameters were not recorded. Additionally, factors such as the presence of myocardial scar tissue or left ventricular hypertrophy, which were not assessed in our retrospective study, can also significantly influence QRSarea. These unmeasured variables could be a source of the observed discrepancy. We postulate that QRSarea may prove more reliable in within-patient comparisons of different pacing positions or device settings, as this would eliminate the variability of R-wave magnitude and other patient-specific factors between patients.

One surprising finding was the unusually low SDAT value observed in a patient with the RV electrode positioned in segment 15 (free wall-mid-inferior), which skewed the segment’s data. Removing this outlier would increase the mean SDAT of that segment to 35 ms, aligning more closely with expected results. A similar anomaly was noted in segment 18 (free wall-basal-inferior), where a patient exhibited a very low SDAT despite having a free wall electrode placement. These deviations emphasize the potential impact of electrode localization inaccuracies and the limitations posed by the relatively small sample size of the study.

### Limitations

A significant limitation of this study lies in its retrospective nature and reliance on EMR data. The imaging modalities, such as ultrasounds and CT scans, were performed for other clinical purposes and were not optimized to pinpoint the exact location of the RV electrode tip, which may have contributed to potential localization inaccuracies. The small sample size (n=65) and the fact that 27 out of 65 patients had incomplete localization data are also notable limitations. Furthermore, our study relied on literature-derived hemodynamic data from Vancura et al. as the ground truth rather than performing direct hemodynamic measurements, which is a key limitation. Although QRSarea might be valuable for intra-patient comparisons, this specific application was not tested in the current study.

The absence of patient demographic data, such as age and sex, also limits our ability to analyze how these factors might influence the relationship between pacing location and dyssynchrony, which may restrict the broad generalizability of our findings to other patient populations.

### Implications and Future Research

The primary objective of this study was hypothesis generation. Establishing a causal relationship between dyssynchrony parameters and hemodynamic outcomes will require direct hemodynamic studies, which fall beyond the scope of this research. Nonetheless, our findings demonstrate that key observations from Vancura et al. can be approximated using vectorcardiography-based SDAT.

Virtual Body Surface Mapping (VBSM) offers numerous advantages for clinical practice. It requires only four electrodes and a lightweight iOS-based device, such as a tablet. The system combines the strengths of 12-lead ECG with vectorcardiography parameters, including QRS area, and body surface mapping parameters such as standard deviation of activation times (SDAT). Additionally, it provides precise visualization of activation times in real time on a beat-to-beat basis. Importantly, VBSM does not interfere with fluoroscopy and can be used intraoperatively without disrupting the sterile field.

If implemented intraoperatively, minimizing VBSM-derived SDAT and other dyssynchrony parameters could guide RV electrode placement, potentially resulting in improved hemodynamic outcomes. The development of the VBSM system used in our study is part of a broader trend toward non-invasive technologies that provide greater insight into cardiac electrical activity. Other emerging methods, such as ultra-high frequency ECG (UHF-ECG), also hold promise for evaluating electrical activation and could offer complementary or alternative tools for clinical decision-making. Future research is essential to directly validate the relationship between vectorcardiography- and VBSM-derived dyssynchrony metrics and hemodynamic performance. Such studies could further refine noninvasive tools for optimizing pacing strategies and enhance clinical outcomes for patients requiring RV pacing.

### Conclusion

In conclusion, this study demonstrates that SDAT is a valuable parameter for distinguishing between septal and non-septal right ventricular pacing positions, with septal placements consistently showing lower SDAT values. These findings underscore the potential of SDAT in guiding electrode placement to optimize hemodynamic outcomes. However, the retrospective nature, small sample size, and reliance on indirect hemodynamic data are important limitations to consider. Further direct hemodynamic studies are essential to validate these observations and refine the clinical application of SDAT in pacing strategies

## Data Availability

All data produced in the present study are available upon reasonable request to the authors

## Declaration of generative AI and AI-assisted technologies in the writing process

*Statement: During the preparation of this work the author(s) used Google Gemini in order to paraphrase certain passages of the text in order to improve readability in a language that is not our native tongue. After using this tool/service, the author(s) reviewed and edited the content as needed and take(s) full responsibility for the content of the publication*

## References

[1] M. Glikson, J.C. Nielsen, M.B. Kronborg, Y. Michowitz, A. Auricchio, I.M. Barbash, J.A. Barrabes, G. Boriani, F. Braunschweig, M. Brignole, H. Burri, A.J.S. Coats, J.C. Deharo, V. Delgado, G.P. Diller, C.W. Israel, A. Keren, R.E. Knops, D. Kotecha, C. Leclercq, B. Merkely, C. Starck, I. Thylen, J.M. Tolosana, F. Leyva, C. Linde, M. Abdelhamid, V. Aboyans, E. Arbelo, R. Asteggiano, G. Barón-Esquivias, J. Bauersachs, M. Biffi, U. Birgersdotter-Green, M.G. Bongiorni, M.A. Borger, J. Celutkienė, M. Cikes, J.C. Daubert, I. Drossart, K. Ellenbogen, P.M. Elliott, L. Fabritz, V. Falk, L. Fauchier, F. Fernández-Aviles, D. Foldager, F. Gadler, P.G.G. De Vinuesa, B. Gorenek, J.M. Guerra, K. Hermann Haugaa, J. Hendriks, T. Kahan, H.A. Katus, A. Konradi, K.C. Koskinas, H. Law, B.S. Lewis, N.J. Linker, M.L. Løchen, J. Lumens, J. Mascherbauer, W. Mullens, K.V. Nagy, E. Prescott, P. Raatikainen, A. Rakisheva, T. Reichlin, R. Pietro Ricci, E. Shlyakhto, M. Sitges, M. Sousa-Uva, R. Sutton, P. Suwalski, J.H. Svendsen, R.M. Touyz, I.C. Van Gelder, K. Vernooy, J. Waltenberger, Z. Whinnett, K.K. Witte, 2021 ESC Guidelines on cardiac pacing and cardiac resynchronization therapy: Developed by the Task Force on cardiac pacing and cardiac resynchronization therapy of the European Society of Cardiology (ESC) With the special contribution of the European Heart Rhythm Association (EHRA), Europace 24 (2022) 71–164. 10.1093/europace/euab232.

[2] B.L. Wilkoff, J.R. Cook, A.E. Epstein, H.L. Greene, A.P. Hallstrom, H. Hsia, S.P. Kutalek, A. Sharma, Dual Chamber and VVI Implantable Defibrillator Trial Investigators, Dual-chamber pacing or ventricular backup pacing in patients with an implantable defibrillator: the Dual Chamber and VVI Implantable Defibrillator (DAVID) Trial., JAMA 288 (2002) 3115–23. 10.1001/jama.288.24.3115.

[3] G.A. Lamas, K.L. Lee, M.O. Sweeney, R. Silverman, A. Leon, R. Yee, R.A. Marinchak, G. Flaker, E. Schron, E.J. Orav, A.S. Hellkamp, S. Greer, J. McAnulty, K. Ellenbogen, F. Ehlert, R.A. Freedman, N.A.M. Estes, A. Greenspon, L. Goldman, Ventricular Pacing or Dual-Chamber Pacing for Sinus-Node Dysfunction, New England Journal of Medicine 346 (2002) 1854–1862. 10.1056/NEJMoa013040.

[4] V. Vančura, D. Wichterle, V. Melenovský, J. Kautzner, Assessment of optimal right ventricular pacing site using invasive measurement of left ventricular systolic and diastolic function, Europace 15 (2013) 1482–1490. 10.1093/europace/eut068.

[5] A. Dias-Frias, R. Costa, A. Campinas, A. Alexandre, D. Sá-Couto, M.J. Sousa, C. Roque, P. Vieira, V. Lagarto, H. Reis, S. Torres, Right Ventricular Septal Versus Apical Pacing: Long-Term Incidence of Heart Failure and Survival, J Cardiovasc Dev Dis 9 (2022). 10.3390/jcdd9120444.

[6] F. Squara, D. Scarlatti, P. Riccini, G. Garret, P. Moceri, E. Ferrari, Individualized Left Anterior Oblique Projection: A Highly Reliable Patient-Tailored Fluoroscopy Criterion for Right Ventricular Lead Positioning, Circ Arrhythm Electrophysiol 11 (2018). 10.1161/CIRCEP.117.006107.

[7] K. Sedova, K. Repin, G. Donin, P. Van Dam, J. Kautzner, Clinical utility of body surface potential mapping in CRT patients, Arrhythm Electrophysiol Rev 10 (2021) 113–119. 10.15420/aer.2021.14.

[8] A.J. Bank, R.M. Gage, A.E. Curtin, K. V. Burns, J.M. Gillberg, S. Ghosh, Body surface activation mapping of electrical dyssynchrony in cardiac resynchronization therapy patients: Potential for optimization, J Electrocardiol 51 (2018) 534–541. 10.1016/j.jelectrocard.2017.12.004.

[9] R.M. Gage, A.E. Curtin, K. V. Burns, S. Ghosh, J.M. Gillberg, A.J. Bank, Changes in electrical dyssynchrony by body surface mapping predict left ventricular remodeling in patients with cardiac resynchronization therapy, Heart Rhythm 14 (2017) 392–399. 10.1016/j.hrthm.2016.11.019.

[10] W. Ben Johnson, P.J. Vatterott, M.A. Peterson, S. Bagwe, R.D. Underwood, A.J. Bank, R.M. Gage, B. Ramza, B.W. Foreman, V. Splett, T. Haddad, J.M. Gillberg, S. Ghosh, Body surface mapping using an ECG belt to characterize electrical heterogeneity for different left ventricular pacing sites during cardiac resynchronization: Relationship with acute hemodynamic improvement, Heart Rhythm 14 (2017) 385–391. 10.1016/j.hrthm.2016.11.017.

[11] A.M.W. van Stipdonk, I. Ter Horst, M. Kloosterman, E.B. Engels, M. Rienstra, H.J.G.M. Crijns, M.A. Vos, I.C. van Gelder, F.W. Prinzen, M. Meine, A.H. Maass, K. Vernooy, QRS Area Is a Strong Determinant of Outcome in Cardiac Resynchronization Therapy., Circ Arrhythm Electrophysiol 11 (2018) e006497. 10.1161/CIRCEP.118.006497.

[12] N. Tokavanich, N. Prasitlumkum, W. Mongkonsritragoon, A. Trongtorsak, W. Cheungpasitporn, R. Chokesuwattanaskul, QRS area as a predictor of cardiac resynchronization therapy response: A systematic review and meta-analysis., Pacing Clin Electrophysiol 45 (2022) 393–400. 10.1111/pace.14441.

[13] J. Bergquist, L. Rupp, B. Zenger, J. Brundage, A. Busatto, R.S. MacLeod, Body Surface Potential Mapping: Contemporary Applications and Future Perspectives, Hearts 2 (2021) 514–542. 10.3390/hearts2040040.

[14] O.A.E. Salden, A.M.W. van Stipdonk, H.M. den Ruijter, M.J. Cramer, M. Kloosterman, M. Rienstra, A.H. Maass, F.W. Prinzen, K. Vernooy, M. Meine, Heart Size Corrected Electrical Dyssynchrony and Its Impact on Sex-Specific Response to Cardiac Resynchronization Therapy, Circ Arrhythm Electrophysiol 14 (2021). 10.1161/CIRCEP.120.008452.

[15] M.A. Ghossein, A.M.W. van Stipdonk, F. Plesinger, M. Kloosterman, P.C. Wouters, O.A.E. Salden, M. Meine, A.H. Maass, F.W. Prinzen, K. Vernooy, Reduction in the QRS area after cardiac resynchronization therapy is associated with survival and echocardiographic response, J Cardiovasc Electrophysiol 32 (2021) 813–822. 10.1111/jce.14910.

[16] O. Okafor, A. Zegard, P. van Dam, B. Stegemann, T. Qiu, H. Marshall, F. Leyva, Changes in QRS Area and QRS Duration After Cardiac Resynchronization Therapy Predict Cardiac Mortality, Heart Failure Hospitalizations, and Ventricular Arrhythmias, J Am Heart Assoc 8 (2019). 10.1161/JAHA.119.013539.

